# Weakly supervised and transfer learning for adenocarcinoma classification in transurethral resection of the prostate whole slide images

**DOI:** 10.1101/2022.04.20.22274062

**Authors:** Masayuki Tsuneki, Makoto Abe, Fahdi Kanavati

## Abstract

The transurethral resection of the prostate (TUR-P) is generally considered an option for benign prostatic diseases especially nodular hyperplasia patients who have moderate to severe urinary problems that have not responded to medication. Importantly, incidental prostate cancer are diagnosed at the time of TUR-P for benign prostatic disease. Since diagnosing a large number of cases containing TUR-P specimens which are characterized by a very large volume of tissue fragments by pathologists using a conventional microscope is time-consuming and limited in terms of human resources. Thus, it is necessary to develop new techniques which can rapidly and accurately screen large numbers of TUR-P specimens. Computational pathology applications which can assist pathologists in detecting prostate adenocarcinoma from TUR-P whole slide images (WSIs) would be of great benefit for routine histopathological workflow. In this study, we trained deep learning models to classify TUR-P WSIs into prostate adenocarcinoma and benign (non-neoplastic) lesions using transfer and weakly supervised learning. We evaluated the models on TUR-P, needle biopsy, and The Cancer Genome Atlas (TCGA) public dataset test sets, achieving an ROC-AUC up to 0.984 in TUR-P test sets for adenocarcinoma. The results demonstrate the high promising potential of deployment in a practical TUR-P histopathological diagnostic workflow system.

## 1 Introduction

According to the Global Cancer Statistics 2020, prostate cancer is the most frequently diagnosed cancer in men in over one-half (112 of 185) of the countries in the world and the second-most-frequent cancer and the fifth leading cause of cancer death among men in 2020 with an estimated 1,414,259 new cases and 375,304 deaths worldwide Sung et al (2021). Histopathological confirmation is necessary because early carcinomas are not able to be distinguished with assurance from foci of nodular hyperplasia and inflammatory lesions.

Nodular hyperplasia (benign prostatic hyperplasia) is a common benign disorder of the prostate which represents a nodular enlargement of the gland caused by hyperplasia of both glandular and stromal components, resulting in an increase in the weight of the prostate. The conventional treatment for nodular hyperplasia is surgical and the transurethral resection of the prostate (TUR-P) is one of the widely practiced surgical procedures, which is estimated to perform about 7,864 cases annually in 2014 in Japan Takamori et al (2017). TUR-P can be both diagnostic and therapeutic when patients have obstructive symptoms, high PSA, and negative prostate needle biopsies Dellavedova et al (2010). In TUR-P, an electrical loop of resectoscope excises hyperplastic prostate tissues to improve urine flow, resulting to produce many tiny tissue fragments during the procedure. Therefore, there are many wide range of tissue fragments in a single slide glass to be observed by pathologists. As compared to conventional biopsy specimens (e.g., endoscopic biopsy from gastrointestinal tracts), TUR-P specimens are characterized by a very large volume of tissues and a large number of glass slides; therefore, histopathological diagnosis for TUR-P specimen is one of the most tedious and error-prone tasks because it is difficult to determine the orientation of the specimen as well as there are strong tissue artifacts. Importantly, cancers especially prostate adenocarcinoma are detected incidentally around 5-17% of TUR-P specimens Jones et al (2009); Zigeuner et al (2003); Yoo et al (2012); Sakamoto et al (2014); Trpkov et al (2008); Dellavedova et al (2010); Otto et al (2014). Conventional active treatment (surgery or radiotherapy) is indicated in T1a patients with life expectancy longer than 10 years, and in the majority of T1b patients Dellavedova et al (2010). Precised histopathological evaluation of cancer (adenocarcinoma) in TUR-P specimen is very important because the presence of cancer in more than 5% of the tissue fragments Epstein et al (1986); Van Andel et al (1995) or high-grade cancer Andrén et al (2006); Egevad et al (2002) may affect the choice of treatment. Thus, for TUR-P specimens, reporting both the number of microscopic foci of carcinoma and the percentage of carcinomatous involvement is recommended. All these factors and burdens mentioned above highlight the benefit of establishing a histopathological screening system to detect prostate adenocarcinoma based on TUR-P specimens. Conventional glass slides of TUR-P specimens can be digitized as whole slide images (WSIs) which could great benefit from the application of computational histopathology algorithms especially deep learning models to aid pathologists allowing the potential of reducing the burden of time-consuming diagnosis and increasing the appropriate detection rate of prostate adenocarcinoma in TUR-P WSIs as part of a screening system.

In computational pathology, deep learning models have been widely applied in histopathological cancer classification on WSIs, cancer cell detection and segmentation, and the stratification of patient outcomes Yu et al (2016); Hou et al (2016); Madabhushi and Lee (2016); Litjens et al (2016); Kraus et al (2016); Korbar et al (2017); Luo et al (2017); Coudray et al (2018); Wei et al (2019); Gertych et al (2019); Bejnordi et al (2017); Saltz et al (2018); Campanella et al (2019); Iizuka et al (2020). Previous works have looked into applying deep learning models for adenocarcinoma classification in stomach Iizuka et al (2020); Kanavati and Tsuneki (2021b); Kanavati et al (2021a), colon Iizuka et al (2020); Tsuneki and Kanavati (2021), lung Kanavati and Tsuneki (2021b); Kanavati et al (2021b), and breast Kanavati and Tsuneki (2021a); Kanavati et al (2022) histopathological specimen WSIs. In a previous study, we trained a prostate adenocarcinoma classification model on needle biopsy WSIs Tsuneki et al (2022) and evaluated the models on both needle biopsy and TUR-P WSI test sets to confirm their application on different types of specimens, achieving an ROC-AUC up to 0.978 in needle biopsy test sets; however, the model under-performed on TUR-P WSIs. Therefore, in this study, we trained deep learning models specifically for TUR-P WSIs. We evaluated the trained models on TUR-P, needle biopsy, and TCGA (The Cancer Genome Atlas) public dataset test sets, achieving an ROC-AUC up to 0.984 in TUR-P test sets, 0.913 in needle biopsy test sets, and 0.947 in TCGA public dataset test sets. These findings suggest that deep learning models might be very useful as routine histopathological diagnostic aids for inspecting TUR-P WSIs to detect prostate adenocarcinoma precisely.

## 2 Materials and methods

### 2.1 Clinical cases and pathological records

Retrospectively, a total of 2,060 H&E (hematoxylin & eosin) stained histopathological specimen slides of human prostate adenocarcinoma and benign (non-neoplastic) lesions –1,560 TUR-P and 500 needle biopsy– were collected from the surgical pathology files of total three hospitals: Shinyukuhashi, Wajiro, and Shinkuki hospitals (Kamachi Group Hospitals, Fukuoka, Japan), after histopathological review of all specimens by surgical pathologists. The histopathological specimens were selected randomly to reflect a real clinical settings as much as possible. Prior to the experimental procedures, each WSI diagnosis was observed by at least two pathologists with the final checking and verification performed by senior pathologists. All WSIs were scanned at a magnification of x20 using the same Leica Aperio AT2 Digital Whole Slide Scanner (Leica Biosystems, Tokyo, Japan) and were saved as SVS file format with JPEG2000 compression.

### 2.2 Dataset

Hospitals which provided histopathological specimen slides in the present study were anonymised (e.g., Hospital-A, B, and C). Table 1 breaks down the distribution of training and validation sets of TUR-P WSIs from two domestic hospitals (Hospital-A and B). Validation sets were selected randomly from the training sets (Table 1). The test sets consisted of TUR-P, needle biopsy, and TCGA (The Cancer Genome Atlas) public dataset WSIs (Table 2). The distribution of test sets from three domestic hospitals (Hospital-A, B, and C) and TCGA public dataset was summarized in Table 2. The patients’ pathological records were used to extract the WSIs’ pathological diagnoses and to assign WSI labels. Training set WSIs were not annotated, and the training algorithm only used the WSI diagnosis labels, meaning that the only information available for the training was whether the WSI contained adenocarcinoma or was benign (non-neoplastic lesion), but no information about the location of the cancerous tissue lesions. The external prostate TCGA datasets are publicly available through the Genomic Data Commons (GDC) Data Portal (https://portal.gdc.cancer.gov/). We have confirmed that surgical pathologists were able to diagnose test sets in Table 2 from visual inspection of the H&E stained slide WSIs alone.

**Table 1:**
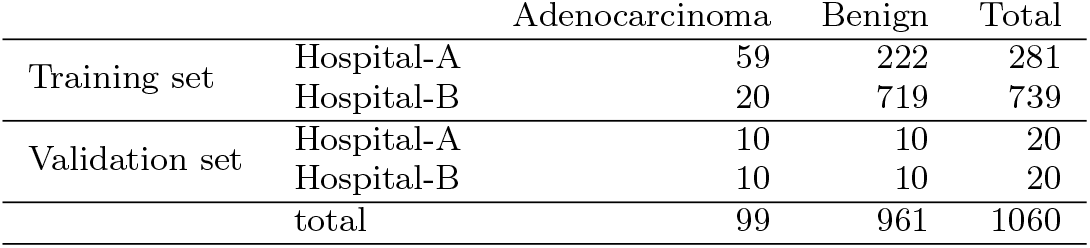
Distribution of transuretheral resection of the prostate (TUR-P) whole-slide images (WSIs) in the training and validation sets obtained from two hospitals (A and B)

**Table 2:** Distribution of whole slide images (WSIs) in the transuretheral resection of the prostate (TUR-P), public dataset (TCGA), and needle biopsy test sets obtained from three hospitals (A-C)

|  |  | Adenocarcinoma | Benign | Total |
| --- | --- | --- | --- | --- |
| TUR-P | Hospital-A-B | 70 | 430 | 500 |
|  | Hospital-A | 40 | 120 | 160 |
|  | Hospital-B | 30 | 310 | 340 |
| Public dataset | TCGA | 733 | 34 | 767 |
| Needle biopsy | Hospital-A-C | 250 | 250 | 500 |

### 2.3 Deep learning models

We trained the models via transfer learning using the partial fine-tuning approachKanavati and Tsuneki (2021c). This is an efficient fine-tuning approach that consists of using the weights of an existing pre-trained model and only fine-tuning the affine parameters of the batch normalization layers and the final classification layer. For the model architecture, we used EfficientNetB1 Tan and Le (2019) starting with pre-trained weights on ImageNet. Figure 1 shows an overview of the training method. The training methodology that we used in the present study was exactly the same as reported in our previous studies Kanavati and Tsuneki (2021b); Tsuneki et al (2022). For completeness we repeat the methodology here.

**Fig. 1:**
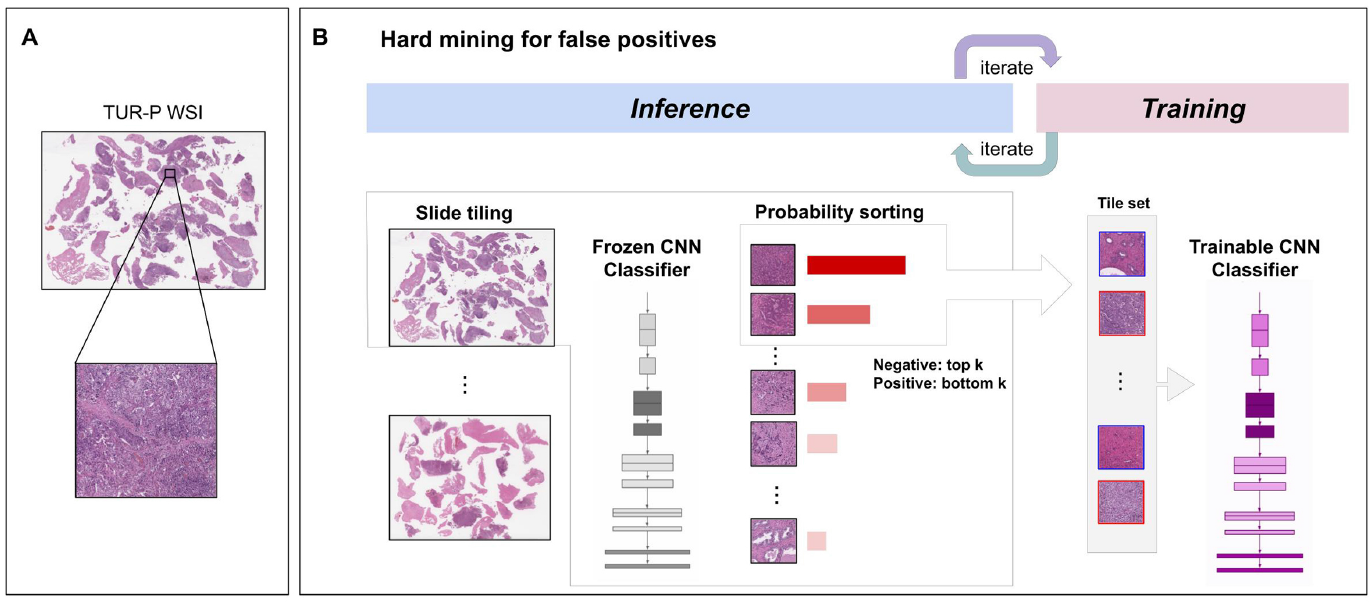
Schematic diagrams of training method overview. (A) shows a representative zoomed-in example of a histopathological patch from a transurethral resection of the prostate (TUR-P) whole slide image (WSI). During training (B), we iteratively alternated between inference and training. During the inference step, the model weights were frozen and the model was used to select tiles with the highest probability after applying it on the entire tissue regions of each WSI. The top k tiles with the highest probabilities were then selected from each WSI and placed into a queue. During training, the selected tiles from multiple WSIs formed a training batch and were used to train the model.

We performed slide tiling by extracting square tiles from tissue regions of the WSIs. We started by detecting the tissue regions in order to eliminate most of the white background. We did this by performing a thresholding on a grayscale version of the WSIs using Otsu’s method Otsu (1979). During prediction, we performed the tiling of the tissue regions in a sliding window fashion, using a fixed-size stride. During training, we initially performed random balanced sampling of tiles extracted from the tissue regions, where we tried to maintain an equal balance of each label in the training batch. To do so, we placed the WSIs in a shuffled queue such that we looped over the labels in succession (i.e., we alternated between picking a WSI with a positive label and a negative label). Once a WSI was selected, we randomly sampled 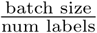 tiles from each WSI to form a balanced batch.

To maintain the balance on the WSI, we oversampled from the WSIs to ensure the model trained on tiles from all of the WSIs in each epoch. We then switched to hard mining of tiles. To perform the hard mining, we alternated between training and inference. During inference, the CNN was applied in a sliding window fashion on all of the tissue regions in the WSI, and we then selected the *k* tiles with the highest probability for being positive. This step effectively selects the tiles that are most likely to be false positives when the WSI is negative. The selected tiles were placed in a training subset, and once that subset contained *N* tiles, the training was run. We used *k* = 8, *N* = 256, and a batch size of 32.

To obtain a single prediction for the WSIs from the the tile predictions, we took the maximum probability from all of the tiles. We used the Adam optimizer Kingma and Ba (2014), with the binary cross-entropy as the loss function, with the following parameters: *beta*_1_ = 0.9, *beta*_2_ = 0.999, a batch size of 32, and a learning rate of 0.001 when fine-tuning. We used early stopping by tracking the performance of the model on a validation set, and training was stopped automatically when there was no further improvement on the validation loss for 10 epochs. We chose the model with the lowest validation loss as the final model.

### 2.4 Software and statistical analysis

The deep learning models were implemented and trained using TensorFlow Abadi et al (2015). AUCs were calculated in python using the scikit-learn package Pedregosa et al (2011) and plotted using matplotlib Hunter (2007). The 95% CIs of the AUCs were estimated using the bootstrap method Efron and Tibshirani (1994) with 1000 iterations.

The true positive rate (TPR) was computed as

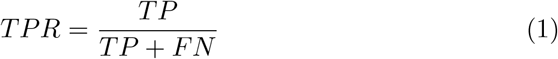

and the false positive rate (FPR) was computed as

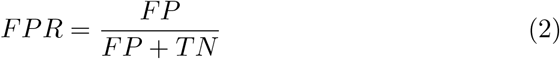

Where TP, FP, and TN represent true positive, false positive, and true negative, respectively. The ROC curve was computed by varying the probability threshold from 0.0 to 1.0 and computing both the TPR and FPR at the given threshold.

### 2.5 Availability of data and material

The datasets generated during and/or analysed during the current study are not publicly available due to specific institutional requirements governing privacy protection but are available from the corresponding author on reasonable request. The datasets that support the findings of this study are available from Kamachi Group Hospitals (Fukuoka, Japan), but restrictions apply to the availability of these data, which were used under a data use agreement which was made according to the Ethical Guidelines for Medical and Health Research Involving Human Subjects as set by the Japanese Ministry of Health, Labour and Welfare (Tokyo, Japan), and so are not publicly available. However, the data are available from the authors upon reasonable request for private viewing and with permission from the corresponding medical institutions within the terms of the data use agreement and if compliant with the ethical and legal requirements as stipulated by the Japanese Ministry of Health, Labour and Welfare.

### 2.6 Code availability

To train the classification model in this study we adapted the publicly available TensorFlow training script available at https://github.com/tensorflow/models/tree/master/official/vision/image_classification.

## 3 Results

### 3.1 Insufficient AUC performance of WSI prostate adenocarcinoma evaluation on TUR-P WSIs using existing series of adenocarcinoma classification models

Prior to the training of prostate adenocarcinoma model using TUR-P WSIs, we have demonstrated the existing adenocarcinoma classification models AUC performances on TUR-P test sets (Table 2). Existing adenocarcinoma classification models were summarized in Table 3: (1) breast invasive ductal carcinoma (IDC) classification model (Breast IDC (x10, 512)) Kanavati and Tsuneki (2021a), (2) breast invasive ductal carcinoma and ductal carcinoma in-situ (DCIS) classification model (Breast IDC, DCIS (x10, 224)) Kanavati et al (2022), (3) colon adenocarcinoma (ADC) and adenoma (AD) classification model (Colon ADC, AD (x10, 512)) Iizuka et al (2020), (4) colon poorly differentiated adenocarcinoma classification model (transfer learning model from stomach poorly differentiated adenocarcinoma classification model) (Colon poorly ADC-1 (x20, 512)) Tsuneki and Kanavati (2021), (5) colon poorly differentiated adenocarcinoma classification model (EfficientNetB1 trained model) (Colon poorly ADC-2 (x20, 512)) Tsuneki and Kanavati (2021), (6) stomach adenocarcinoma and adenoma classification model (Stomach ADC, AD (x10, 512)) Iizuka et al (2020), (7) stomach poorly differentiated adenocarcinoma classification model (Stomach poorly ADC (x20, 224)) Kanavati and Tsuneki (2021b), (8) stomach signet ring cell carcinoma (SRCC) classification model (Stomach SRCC (x10, 224)) Kanavati et al (2021a), (9) pancreas endoscopic ultrasound guided fine needle aspiration (EUS-FNA) biopsy adenocarcinoma classification model (Pancreas EUS-FNA ADC (x10, 224)) Naito et al (2021), and (10) lung carcinoma classification model (Lung Carcinoma (x10, 512)) Kanavati et al (2020). Table 3 shows that Colon poorly ADC-2 (x20, 512) and Lung Carcinoma (x10, 512) models exhibited both high ROC-AUC and low log loss values as compared to other models. Thus, we have trained the models based on the Colon poorly ADC-2 (x20, 512) and Lung Carcinoma (x10, 512) models using TUR-P training sets (Table 1).

**Table 3:** ROC-AUC and log loss results for adenocarcinoma classification on transuretheral resection of the prostate (TUR-P) test sets (Hospital-A-B) using existing adenocarcinoma classification models

| Existing models | ROC-AUC | Log loss |
| --- | --- | --- |
| Breast IDC (x10, 512) | 0.737 [0.664 - 0.807] | 1.428 [1.340 - 1.530] |
| Breast IDC, DCIS (x10, 224) | 0.635 [0.565 - 0.720] | 3.783 [3.624 - 3.929] |
| Colon ADC, AD (x10, 512) | 0.608 [0.546 - 0.679] | 3.812 [3.595 - 4.028] |
| Colon poorly ADC-1 (x20, 512) | 0.780 [0.713 - 0.840] | 0.863 [0.811 - 0.913] |
| Colon poorly ADC-2 (x20, 512) | 0.771 [0.681 - 0.837] | 0.859 [0.890 - 0.914] |
| Stomach ADC, AD (x10, 512) | 0.762 [0.689 - 0.833] | 3.133 [2.948 - 3.268] |
| Stomach poorly ADC (x20, 224) | 0.617 [0.529 - 0.698] | 1.588 [1.504 - 1.657] |
| Stomach SRCC (x10, 224) | 0.670 [0.600 - 0.734] | 0.549 [0.499 - 0.606] |
| Pancreas EUS-FNA ADC (x10, 224) | 0.808 [0.746 - 0.888] | 1.080 [1.031 - 1.142] |
| Lung Carcinoma (x10, 512) | 0.737 [0.662 - 0.801] | 0.357 [0.298 - 0.423] |

### 3.2 High AUC performance of TUR-P WSI evaluation of prostate adenocarcinoma histopathology images

We have trained models using transfer learning (TL) and weakly supervised learning approaches which could be used with weak labels (WSI labels) Kanavati et al (2020); Tsuneki et al (2022). At the same time, we trained using the EfficientNetB1 convolutional neural network (CNN) architecture at magnification x10 and x20. The models were applied in a sliding window fashion with input tiles of 224×224 and 512×512 pixels and a stride of 256 (Fig. 1). To train the deep learning models, we used a total of 79 adenocarcinoma and 941 benign training set WSIs and 20 adenocarcinoma and 20 benign validation set WSIs (Table 1). This resulted in four different models: (1) TL-colon poorly ADC-2 (x20, 512), (2) TL-lung carcinoma (x10, 512), (3) EfficientNetB1 (x10, 224), and (4) EfficientNetB1 (x20, 512). We have evaluated four different trained deep learning models on test sets from three different hospitals (Hospital-A-C) and TCGA public datasets (Table 2). For each test set (TUR-P: Hospital-A-B, TUR-P: Hospital-A, TUR-P: Hospital-B, public dataset: TCGA, and needle biopsy: Hospital-A-C), we computed the ROC-AUC, log loss, accuracy, sensitivity, and specificity and summarized in Table 4 & 5 and Fig. 2. The transfer learning model (TL-colon poorly ADC-2 (x20, 512)) (Fig. 2A) from existing colon poorly differentiated adenocarcinoma classification model (Colon poorly ADC-2 (x20, 512)) Tsuneki and Kanavati (2021) trained using TUR-P training sets have higher ROC-AUCs and lower log losses compared to the other models (TL-lung carcinoma (x10, 512), EfficientNetB1 (x10, 224), and EfficientNetB1 (x20, 512)) (Table 4, Fig. 2). On the other hand, on the TUR-P hospital-B test sets, both EfficientNetB1 (x10, 224) and EfficientNetB1 (x20, 512) models exhibited very high ROC-AUCs (0.924 - 0.973) and low log-losses (0.126 - 0.251) as compared to the other test sets (TUR-P Hospital-A, public dataset, and needle biopsy) (Table 4 and Fig. 2C, D). Looking at heatmap images of the same TUR-P WSI which were correctly predicted as prostate adenocarcinoma using four different trained models, both EfficientNetB1 (x10, 224) and EfficientNetB1 (x20, 512) models falsely predicted adenocarcinoma on the marked blue-dots which pathologists marked when they performed diagnosis (Fig. 3C, D). In contrast, both TL-colon poorly ADC-2 (x20, 512) and TL-lung carcinoma (x10, 512) models precisely predicted adenocarcinoma (Fig. 3A, B). The model (TL-colon poorly ADC-2 (x20, 512)) achieved highest ROC-AUC of 0.984 (CI: 0.956 - 1.000) and lowest log loss of 0.127 (CI: 0.076 - 0.205) for prostate adenocarcinoma classification in TUR-P hospital-A test sets and also achieved high ROC-AUCs in public dataset (0.947, CI: 0.922 - 0.972) and needle biopsy test sets (0.913, CI: 0.887 - 0.939) (Table 4). In all test sets, the model ((TL-colon poorly ADC-2 (x20, 512)) achieved very high accuracy (0.821 - 0.969), sensitivity (0.764 - 0.900), and specificity (0.884 - 0.992) (Table 5). As shown in Fig. 2 & 3 and Table 4 & 5, the model (TL-colon poorly ADC-2 (x20, 512)) is fully applicable for prostate adenocarcinoma classification in TUR-P WSIs as well as TCGA public WSI dataset and even needle biopsy WSIs. Figures 4, 5, 6, 7, and 8 show representative WSIs of true-positive, true-negative, false-positive, and false-negative, respectively from using the model (TL-colon poorly ADC-2 (x20, 512)).

**Table 4:** ROC-AUC and log loss results for adenocarcinoma classification on the transuretheral resection of the prostate (TUR-P), public dataset (TCGA), and needle biopsy test sets using trained models

|  |  | TL-colon poorly ADC-2 (x20, 512) |  |
| --- | --- | --- | --- |
|  |  | ROC-AUC | Log-loss |
| TUR-P | Hospital-A-B | 0.947 [0.910 - 0.976] | 0.191 [0.146 - 0.242] |
|  | Hospital-A | 0.984 [0.956 - 1.000] | 0.127 [0.076 - 0.205] |
|  | Hospital-B | 0.896 [0.822 - 0.956] | 0.221 [0.160 - 0.299] |
| Public dataset | TCGA | 0.947 [0.922 - 0.972] | 0.335 [0.288 - 0.390] |
| Needle biopsy | Hospital-A-C | 0.913 [0.887 - 0.939] | 0.587 [0.480 - 0.700] |

|  |  | TL-lung carcinoma (x10, 512) |  |
| --- | --- | --- | --- |
|  |  | ROC-AUC | Log-loss |
| TUR-P | Hospital-A-B | 0.892 [0.860 - 0.948] | 0.328 [0.282 - 0.364] |
|  | Hospital-A | 0.972 [0.917 - 0.998] | 0.277 [0.217 - 0.364] |
|  | Hospital-B | 0.785 [0.688 - 0.870] | 0.351 [0.301 - 0.403] |
| Public dataset | TCGA | 0.878 [0.822 - 0.929] | 0.258 [0.213 - 0.299] |
| Needle biopsy | Hospital-A-C | 0.826 [0.786 - 0.860] | 0.808 [0.702 - 0.931] |

|  |  | EfficientNetB1 (x10, 224) |  |
| --- | --- | --- | --- |
|  |  | ROC-AUC | Log-loss |
| TUR-P | Hospital-A-B | 0.885 [0.829 - 0.927] | 0.239 [0.181 - 0.298] |
|  | Hospital-A | 0.837 [0.752 - 0.909] | 0.479 [0.318 - 0.619] |
|  | Hospital-B | 0.973 [0.916 - 1.000] | 0.126 [0.092 - 0.168] |
| Public dataset | TCGA | 0.639 [0.563 - 0.716] | 3.800 [3.613 - 3.977] |
| Needle biopsy | Hospital-A-C | 0.779 [0.736 - 0.822] | 0.659 [0.552 - 0.769] |

**Table 4:** ROC-AUC and log loss results for adenocarcinoma classification on the transurethral resection of the prostate (TUR-P), public dataset (TCGA), and needle biopsy test sets using trained models
|  |  | EfficientNetB1 (x20, 512) |  |
| --- | --- | --- | --- |
|  |  | ROC-AUC | Log-loss |
| TUR-P | Hospital-A-B | 0.840 [0.767 - 0.897] | 0.315 [0.269 - 0.377] |
|  | Hospital-A | 0.794 [0.681 - 0.872] | 0.451 [0.323 - 0.601] |
|  | Hospital-B | 0.924 [0.856 - 0.998] | 0.251 [0.203 - 0.290] |
| Public dataset | TCGA | 0.533 [0.464 - 0.616] | 2.611 [2.485 - 2.721] |
| Needle biopsy | Hospital-A-C | 0.655 [0.609 - 0.702] | 1.785 [1.551 - 1.956] |

**Table 5:** Scores of accuracy, sensitivity, and specificity on the transuretheral resection of the prostate (TUR-P), public dataset (TCGA), and needle biopsy test sets using the best model (TL-colon poorly ADC-2 (x20, 512))

|  |  | Accuracy | Sensitivity | Specificity |
| --- | --- | --- | --- | --- |
| TUR-P | Hospital-A-B | 0.916 [0.892 - 0.938] | 0.871 [0.794 - 0.951] | 0.923 [0.897 - 0.945] |
|  | Hospital-A | 0.969 [0.938 - 0.994] | 0.900 [0.800 - 0.976] | 0.992 [0.974 - 1.000] |
|  | Hospital-B | 0.874 [0.841 - 0.909] | 0.767 [0.613 - 0.906] | 0.884 [0.848 - 0.922] |
| Public dataset | TCGA | 0.821 [0.793 - 0.849] | 0.816 [0.786 - 0.843] | 0.941 [0.852 - 1.000] |
| Needle biopsy | Hospital-A-C | 0.844 [0.812 - 0.874] | 0.764 [0.710 - 0.813] | 0.924 [0.886 - 0.956] |

**Fig. 2:**
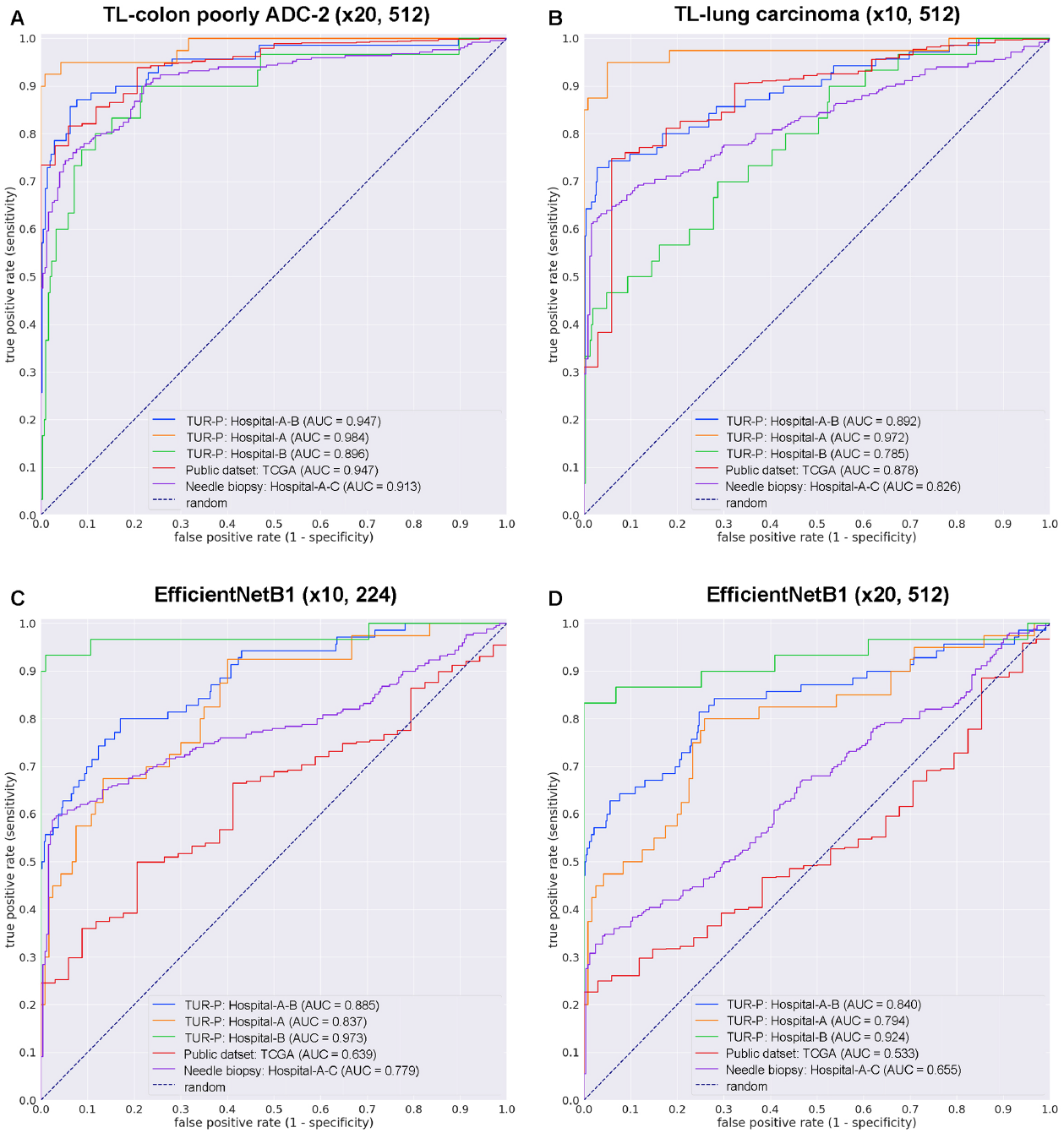
ROC curves with AUCs from four different trained deep learning models (A-D) on the test sets: (A) transfer learning (TL) model from existing colon poorly differentiated adenocarcinoma (ADC) classification model with tile size 224 px and magnification at x20, (B) TL model from existing lung carcinoma classification model with tile size 512 px and magnification at x10, (C) EfficientNetB1 model with tile size 224 px and magnification at x10, and (D) EfficientNetB1 model with tile size 512 px and magnification at x20.

**Fig. 3:**
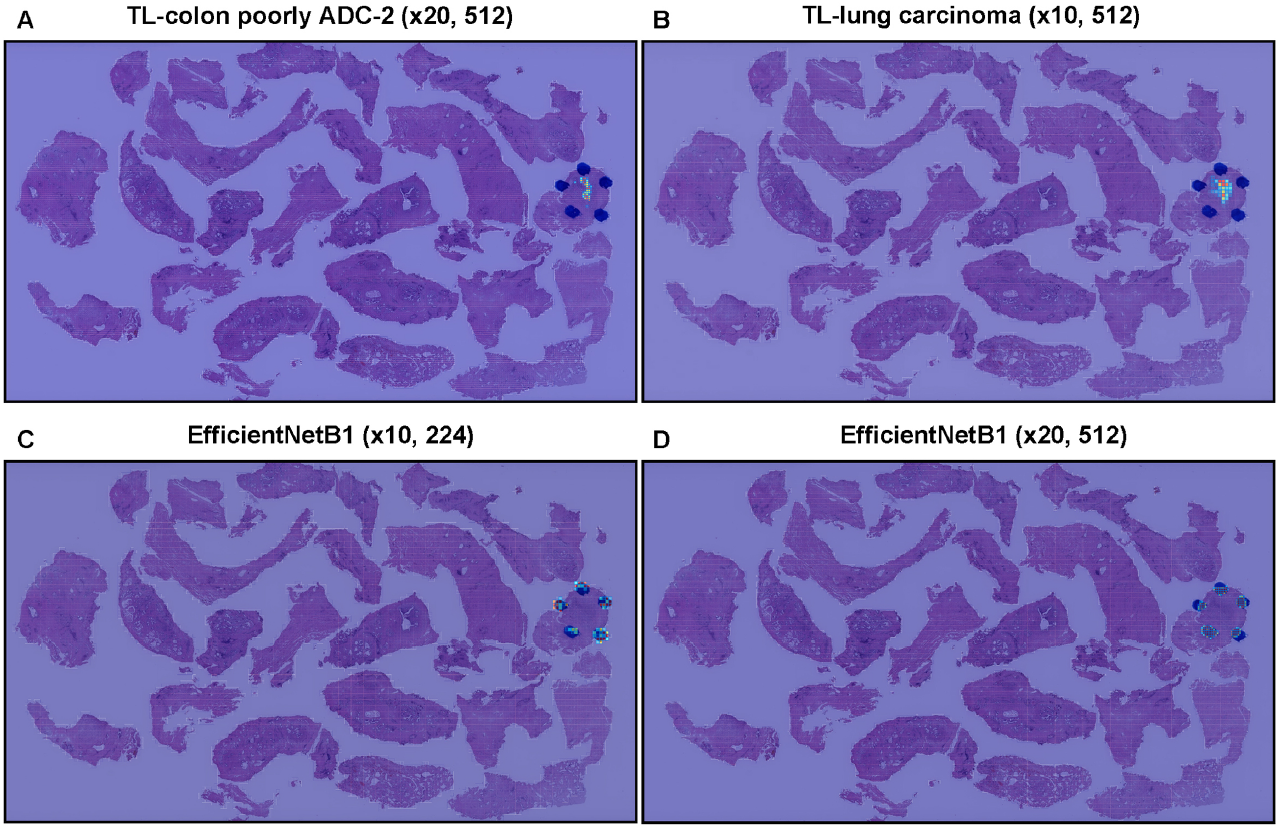
Comparison of adenocarcinoma prediction in transurethral resection of the prostate (TUR-P) whole slide image (WSI) of four trained deep learning models (A-D). In transfer learning (TL) models from colon poorly differentiated adenocarcinoma (A) and lung carcinoma (B), the heatmap images show true positive prediction of adenocarcinoma where pathologists marked surrounding with blue-dots. In EfficientNetB1 models (C, D), the heatmap images show false-positive prediction of adenocarcinoma on the marked blue-dots. The heatmap uses the jet color map where blue indicates low probability and red indicates high probability.

**Fig. 4:**
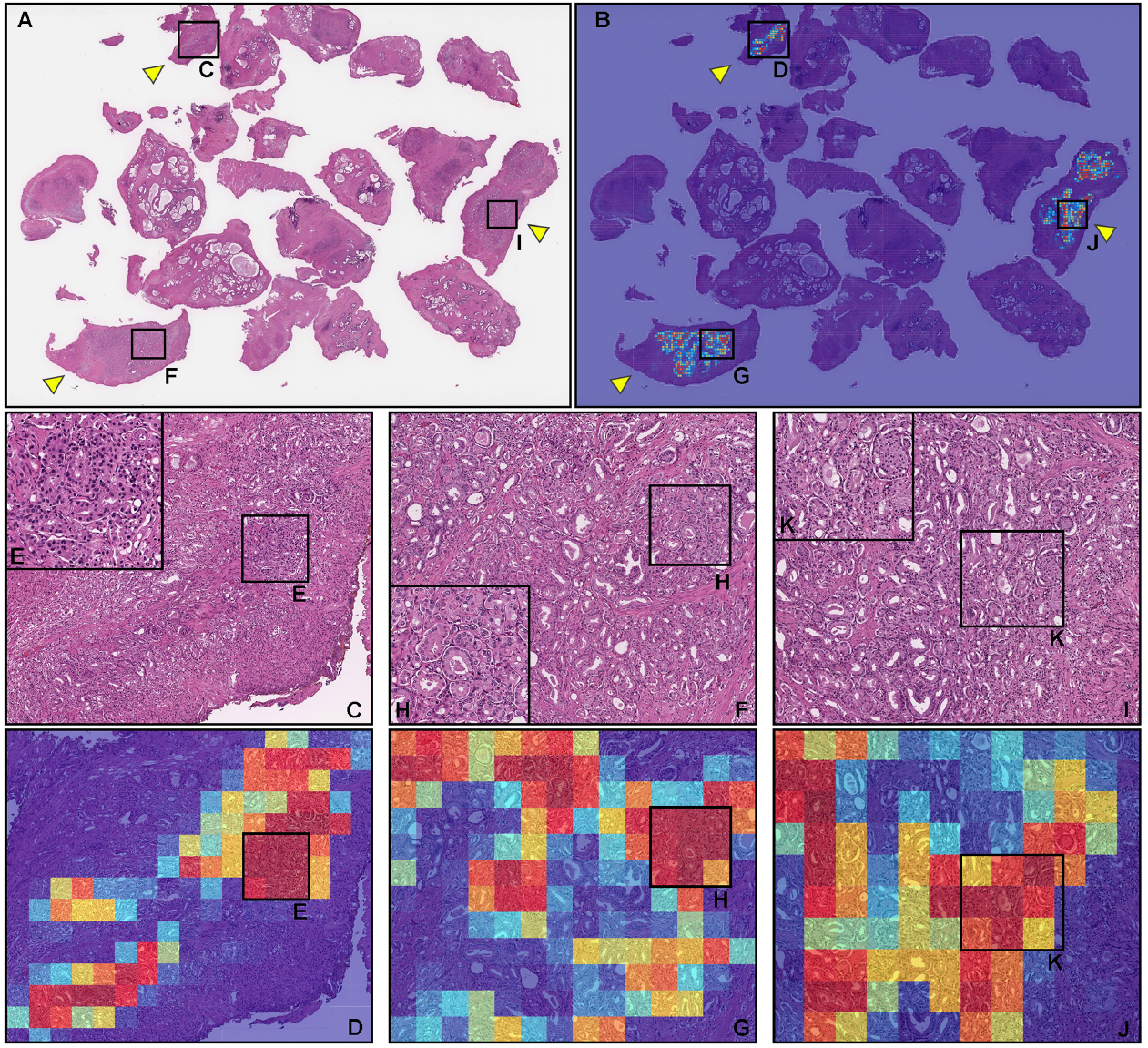
A representative example of prostate adenocarcinoma true positive prediction outputs on a whole slide image (WSI) from transurethral resection of the prostate (TUR-P) test sets using the model (TL-colon poorly ADC-2 (x20, 512)). In the prostate adenocarcinoma WSI of TUR-P specimen (A), according to the histopathological diagnostic report, adenocarcinoma cells infiltrated in the three tissue fragments highlighted with yellow-triangles. The heatmap image (B) shows true positive predictions of prostate adenocarcinoma cells (D, G, J) which correspond respectively to H&E histopathology (C-E, F-H, and I-K). The heatmap image (B) also shows no positive predictions (true negative predictions) in the tissue fragments without evidence of adenocarcinoma infiltration (A). The heatmap uses the jet color map where blue indicates low probability and red indicates high probability.

**Fig. 5:**
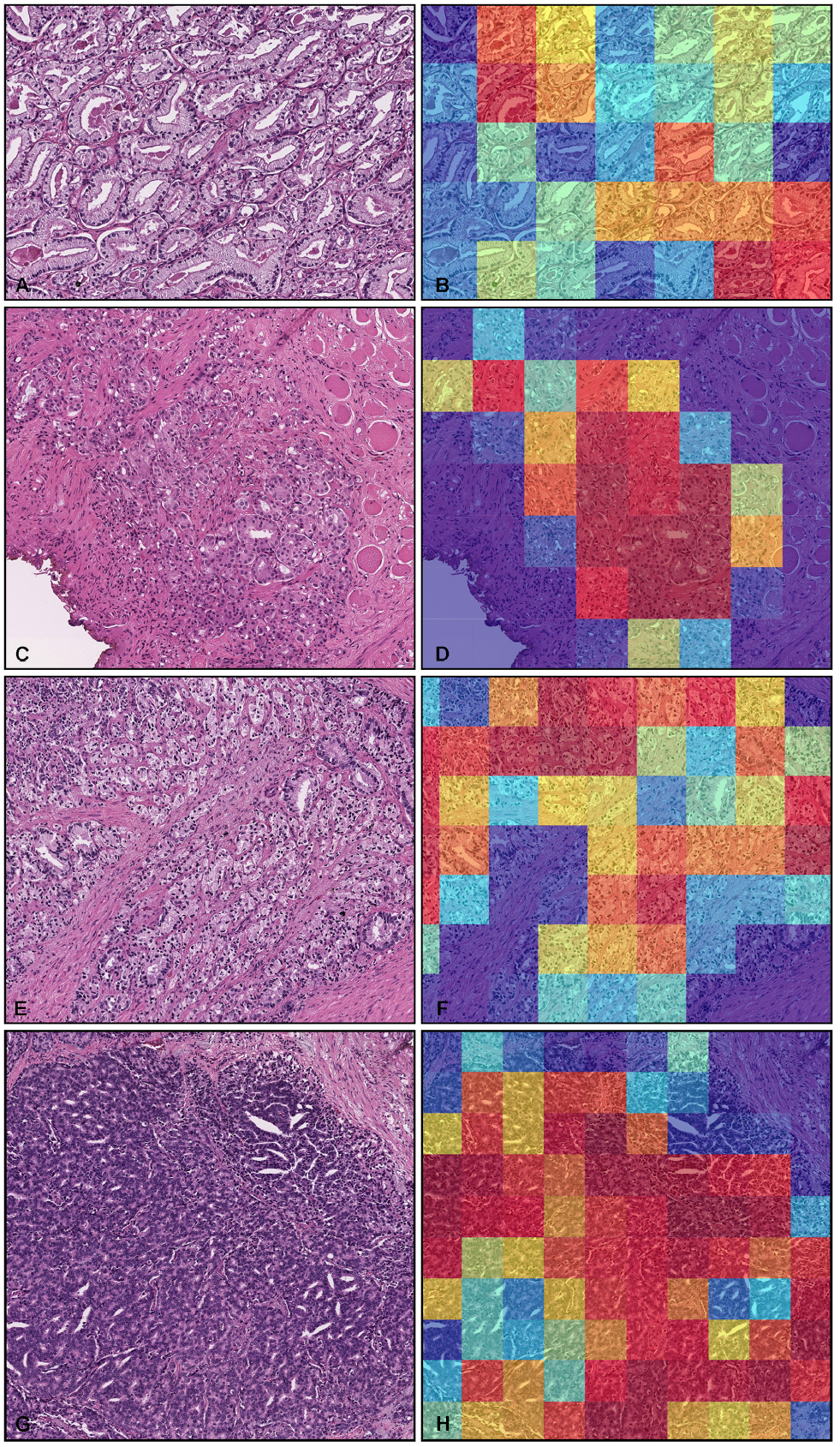
Representative histopathological examples of prostate adenocarcinoma true positive prediction outputs on whole slide images (WSIs) from transurethral resection of the prostate (TUR-P) test sets using the model (TL-colon poorly ADC-2 (x20, 512)). Depiction of prostate adenocarcinoma histopathologies and corresponding heatmap images of adenocarcinoma prediction outputs: (A, B) medium-sized, discrete and distinct neoplastic glands (Gleason pattern 3), (C, D) medium-sized discrete and distinct glands with illformed glands (Gleason score 3+4), (E, F) ill-formed glands (Gleason pattern 4), (G, H) cribriform pattern (Gleason pattern 4). The heatmap uses the jet color map where blue indicates low probability and red indicates high probability.

**Fig. 6:**
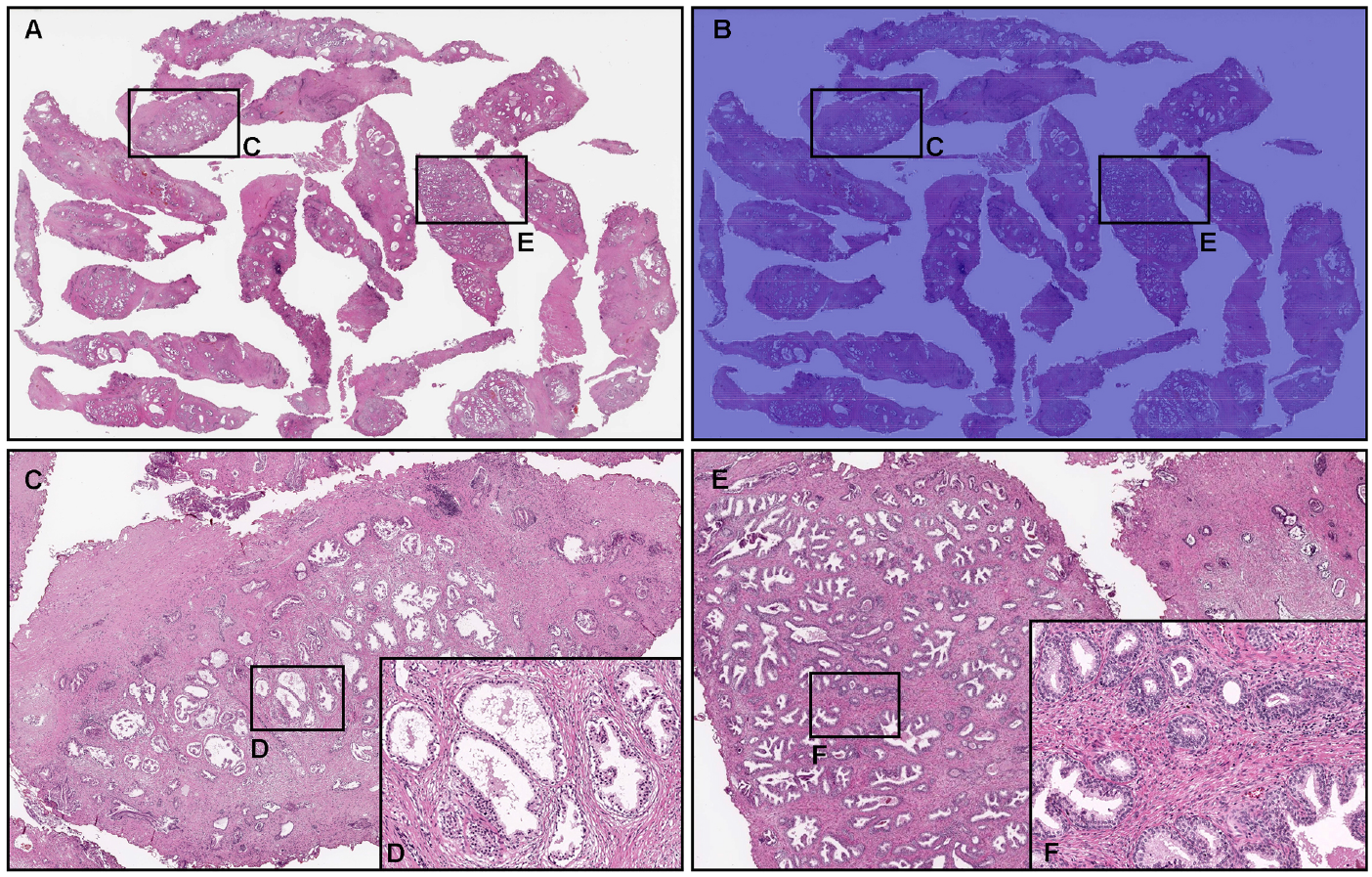
Representative true negative prostate adenocarcinoma prediction outputs on a whole slide image (WSI) from transurethral resection of the prostate (TUR-P) test sets using the model (TL-colon poorly ADC-2 (x20, 512)). Histopathologically, in (A), there were nodular hyperplasia (benign prostatic hyperplasia) with chronic inflammation without any evidence of malignancy (C-E). The heatmap image (B, C, E) shows true negative prediction of prostate adenocarcinoma. The heatmap uses the jet color map where blue indicates low probability and red indicates high probability.

**Fig. 7:**
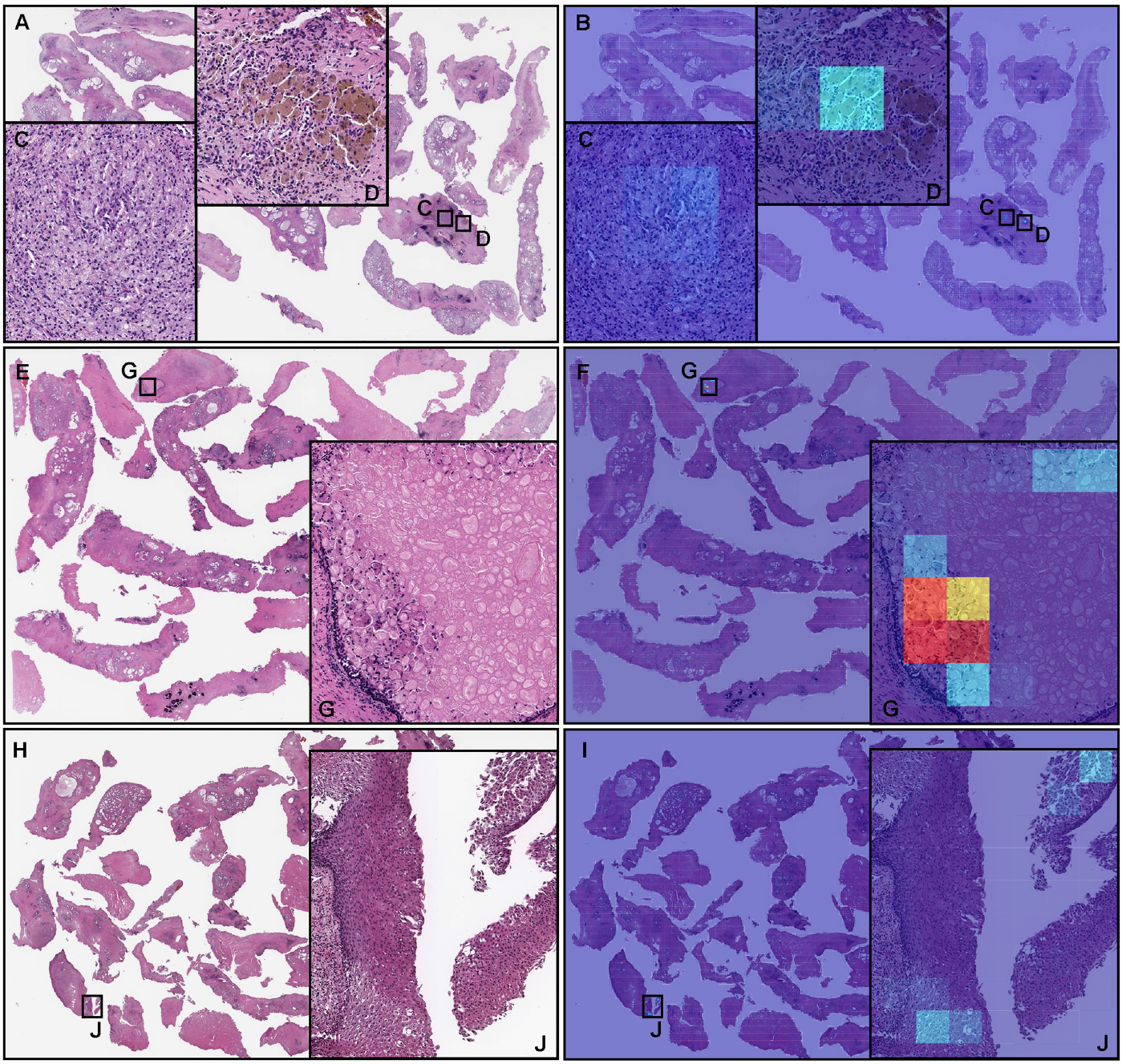
Representative examples of prostate adenocarcinoma false positive prediction outputs on whole slide images (WSIs) from transurethral resection of the prostate (TUR-P) test sets using the model (TL-colon poorly ADC-2 (x20, 512)). Histopathologically, (A, E, and H) have no evidence of adenocarcinoma infiltration. The heatmap images (B, F, and I) exhibit false positive predictions of prostate adenocarcinoma (C, D, G, and J) where the tissues consist of xanthogranulomatous inflammation (C, D), macrophagic infiltration (G), and squamous metaplasia with pseudo-koilocytosis (J), which are most likely the primary cause of the false positive prediction due to its morphological similarity to prostate adenocarcinoma cells. The heatmap uses the jet color map where blue indicates low probability and red indicates high probability.

**Fig. 8:**
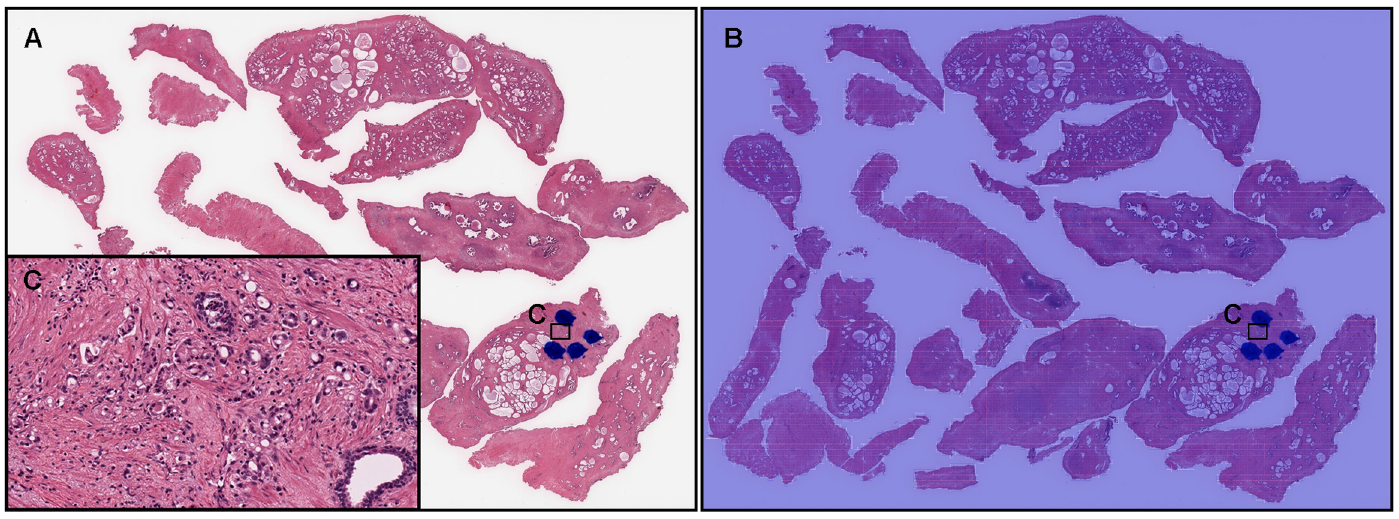
A representative example of prostate adenocarcinoma false negative prediction output on a whole slide image (WSI) from transurethral resection of the prostate (TUR-P) test sets using the model (TL-colon poorly ADC-2 (x20, 512)). According to the histopathological diagnostic report, this case (A) has very small number of adenocarcinoma foci (cells) in (C) where pathologists marked surrounding with blue-dots but not in other areas which consist of nodular hyperplasia. The heatmap image (B) exhibited no positive adenocarcinoma prediction (C). The heatmap uses the jet color map where blue indicates low probability and red indicates high probability.

### 3.3 True positive prostate adenocarcinoma prediction of TUR-P WSIs

Our model (TL-colon poorly ADC-2 (x20, 512)) satisfactorily predicted adenocarcinoma in TUR-P WSIs (Fig. 4A, B). According to the histopathological report and additional pathologist’s reviewing, in this WSI (Fig. 4A), there were three tissue fragments (highlighted with yellow-triangles) with prostate adenocarcinoma cell infiltration (Fig. 4C, E, F, H, I, K). The heatmap image (Fig. 4B) shows true positive predictions in these fragments (yellow-triangles) (Fig. 4D, E, G, H, J, K) without false-positive predictions in other tissue fragments which were histopathologically evaluated as nodular hyperplasia (benign prostatic hyperplasia) without evidence of malignancy (Fig. 4A, B). Not only this representative WSI (Fig. 4), our model (TL-colon poorly ADC-2 (x20, 512)) precisely predicted wide variety of prostate adenocarcinoma histopathological features (Fig. 5): medium-sized, discrete and distinct neoplastic glands (Gleason pattern 3) (Fig. 5A, B); medium-sized discrete and distinct glands with ill-formed glands (Gleason score 3+4) (Fig. 5C, D); ill-formed glands (Gleason pattern 4) (Fig. 5E, F); and cribriform pattern (Gleason pattern 4) (Fig. 5G, H).

### 3.4 True negative prostate adenocarcinoma prediction of TUR-P WSIs

Our model (TL-colon poorly ADC-2 (x20, 512)) showed true negative predictions of prostate adenocarcinoma in TUR-P WSIs (Fig. 6A, B). In Fig. 6A, histopathologically, there were nodular hyperplasia (benign prostatic hyperplasia) with chronic inflammation in all tissue fragments without evidence of malignancy (Fig. 6A, C-F) which were not predicted as prostate adenocarcinoma (Fig. 6B, C, E).

### 3.5 False positive prostate adenocarcinoma prediction of TUR-P WSIs

According to the histopathological reports and additional pathologist’s reviewing, there were no prostate adenocarcinoma in these TUR-P WSIs (Fig. 7A, E, H). Our model (TL-colon poorly ADC-2 (x20, 512)) showed false positive predictions of prostate adenocarcinoma (Fig. 7B-D, F-G, I-J). These false positive tissue areas (Fig. 7B-D, F-G, I-J) showed xanthogranulomatous inflammation (Fig. 7A, C, D), macrophagic infiltration (Fig. 7E, G), and squamous metaplasia with pseudo-koilocytosis (Fig. 7H, J), which could be the primary cause of false positives due to its morphological similarity in adenocarcinoma cells.

### 3.6 False negative prostate adenocarcinoma prediction of TUR-P WSIs

According to the histopathological report and additional pathologist’s reviewing, in this TUR-P WSI (Fig. 8A), there were very small number of adenocarcinoma cells infiltrating in a tissue fragment (Fig. 8C) where pathologists marked with blue-dots. However, our model (TL-colon poorly ADC-2 (x20, 512)) did not predict any prostate adenocarcinoma cells (Fig. 8B, C).

## 4 Discussion

In this study, we trained deep learning models for the classification of prostate adenocarcinoma in TUR-P WSIs. Of the four models we trained (Table 4), the best model (TL-colon poorly ADC-2 (x20, 512)) achieved ROC-AUCs in the range of 0.896 - 0.984 on the TUR-P test sets. The best model (TL-colon poorly ADC-2 (x20, 512)) also achieved high ROC-AUCs on needle biopsy (0.913) and TCGA public dataset (0.947) test sets. The model (TL-lung carcinoma (x10, 512)) also achieved high ROC-AUCs in all test sets but lower than the best one (TL-colon poorly ADC-2 (x20, 512)). The other two models were trained using the EfficientNetB1 Tan and Le (2019) models starting with pre-trained weights on ImageNet at different magnifications (x10 and x20) and tile sizes (224×224 px, 512×512 px). The models based on EfficientNetB1 (EfficientNetB1 (x10, 224) & EfficientNetB1 (x20, 512)) achieved robust high ROC-AUC values on TUR-P Hospital-B test sets as compared to other test sets (Table 4). Based on the prediction heatmap images of prostate adenocarcinoma, it was obvious that the models based on EfficientNetB1 (EfficientNetB1 (x10, 224) & EfficientNetB1 (x20, 512)) incorrectly predicted blue ink dots, which pathologists had marked during diagnosis, as prostate adenocarcinoma (Fig. 3). Based on this finding, we have looked over WSIs in TUR-P Hospital-B test sets (Table 2) and most of adenocarcinoma positive WSIs (28 out of 30 WSIs) had ink dots on WSIs which were falsely predicted as adenocarcinoma. On the other hand, transfer learning models (TL-colon poorly ADC-2 (x20, 512) & TL-lung carcinoma (x10, 512)) revealed no false positive predictions on ink dots (Fig. 3); this is because those models had been trained on WSIs with ink labelled as non-neoplastic. The best model (TL-colon poorly ADC-2 (x20, 512)) and the second best model (TL-lung carcinoma (x10, 512)) were trained by the transfer learning approach from our existing colon poorly differentiated adenocarcinoma classification model Tsuneki and Kanavati (2021) and lung carcinoma classification model Kanavati et al (2020) based on the findings of ROC-AUC and log loss values on TUR-P test sets (TUR-P Hospital-A-B) using existing adenocarcinoma classification models (Table 3). We used the partial fine-tuning approach Kanavati and Tsuneki (2021c) to train the models faster, as there are less weights involved to tune. We used only 1,020 TUR-P WSIs (adenocarcinoma: 79 WSIs, benign: 941 WSIs) (Table 1) without manual annotations by pathologists Iizuka et al (2020); Naito et al (2021); Kanavati et al (2022). We see that by specifically training on TUR-P WSIs, the models significantly improved prediction performance on TUR-P test set (Table 4) compared to the previous study Tsuneki et al (2022) that had lower ROC-AUC (0.737 - 0.909) and higher log loss (3.269 - 4.672) values. The combination of both models would be able to provide accurate prostate adenocarcinoma classification on both needle biopsy Tsuneki et al (2022) and TUR-P WSIs in routine histopathological diagnostic workflow.

Nodular hyperplasia (benign prostatic hyperplasia) is a common benign disorder of the prostate as a histopathological diagnosis referring to the nodular enlargement of the gland caused by hyperplasia of both glandular and stromal components within the prostatic transition zone and results in varying degrees of urinary obstruction, sometimes requiring surgical intervention including TUR-P Lokeshwar et al (2019). Importantly, incidental prostate cancers are diagnosed at the time of TUR-P for benign prostatic disease Otto et al (2014). According to the literature search, cancers especially prostate adenocarcinoma are detected incidentally around 5-17% of TUR-P specimens Jones et al (2009); Zigeuner et al (2003); Yoo et al (2012); Sakamoto et al (2014); Trpkov et al (2008); Dellavedova et al (2010); Otto et al (2014), meaning around 83-95% of TUR-P specimens are benign lesions; which is nearly identical to the ratio of adenocarcinoma in the TUR-P test sets (Table 2). Therefore, the high values of specificity (0.884 - 0.992) in the best model is noteworthy (Table 5). Moreover, heatmap images revealed true-negative prediction perfectly on each non-neoplastic fragment in both adenocarcinoma (Fig. 4) and benign (non-neoplastic) (Fig. 6) WSIs. Thus, the heatmap images predicted by the best model would provide great benefits for pathologists who have to report the detail descriptions of many TUR-P specimens in routine clinical practices.

The best deep learning model established in the present study offers promising results that indicate it could be beneficial as a screening aid for pathologists prior to observing histopathology on glass slides or WSIs. At the same time, the model could be used as a double-check tool to reduce the risk of missed cancer foci (incidental adenocarcinoma in TUR-P specimens). The most important advantage of using a fully automated computational tool is that it can systematically handle large amounts of WSIs without potential bias due to the fatigue commonly experienced by pathologists, which could drastically alleviate the heavy clinical burden of practical pathology diagnosis using conventional microscopes.

## 5 Acknowledgements

We are grateful for the support provided by Dr. Shigeo Nakano at Kamachi Group Hospitals (Fukuoka, Japan). We thank pathologists who have been engaged in reviewing cases and clinicopathological discussion for this study.

## 6 Compliance with Ethical Standards

The experimental protocol was approved by the ethical board of Kamachi Group Hospitals (No. 173). All research activities complied with all relevant ethical regulations and were performed in accordance with relevant guidelines and regulations in the all hospitals mentioned above. Informed consent to use histopathological samples and pathological diagnostic reports for research purposes had previously been obtained from all patients prior to the surgical procedures at all hospitals, and the opportunity for refusal to participate in research had been guaranteed by an opt-out manner.

## 7 Funding

The authors received no financial supports for the research, authorship, and publication of this study.

## 8 Conflict of Interest

M.T. and F.K. are employees of Medmain Inc. All authors declare no competing interests.

## 9 Contributions

M.T. designed the studies, performed experiments and analyzed the data; M.T. and F.K. performed the computational studies; M.A. performed the histopathological diagnoses and reviewed the cases; M.T. and F.K. wrote the manuscript; M.T. supervised the project. All authors reviewed and approved the final manuscript.

